# Association Between Number of Missing Teeth and Abnormal Bowel Habits: A Cross-Sectional Study from NHANES 2005–2010

**DOI:** 10.64898/2026.07.28.26359160

**Authors:** Hao Chen, Jianbo Zhang

## Abstract

**Background/Objectives:** Tooth loss may impair masticatory function, potentially affecting gastrointestinal health. This study examined the association between clinically examined tooth loss and abnormal bowel habits among US adults.

**Methods:** This cross-sectional study used NHANES 2005–2010 data. Adults aged ≥20 with clinical oral examination (OHX) and Bristol Stool Scale (BSS) data were included. Missing teeth (0–28) were categorized as 0, 1–5, 6–10, 11–27, and 28. The primary outcome was abnormal bowel habits (BSS 1–2 or 6–7 vs. 3–5); secondary outcomes included constipation, diarrhea, and fecal incontinence symptoms. Survey-weighted logistic regression with progressive covariate adjustment was used, with Benjamini–Hochberg FDR correction for secondary outcomes.

**Results:** Among 9,988 participants (14.5% prevalence), the P-for-trend for abnormal bowel habits was significant (OR = 1.015, 95% CI: 1.006–1.024, P = 0.001), corroborated by omnibus Wald (P = 0.006) and restricted cubic spline analyses (P = 0.019). Complete edentulism was associated with 1.5-fold higher odds (OR = 1.545, P = 0.006). Constipation survived FDR correction (FDR-adjusted P = 0.032). Results were robust across most of five sensitivity analyses, with modest attenuation noted after denture use adjustment in a restricted subset.

**Conclusions:** Tooth loss was significantly associated with abnormal bowel habits, with constipation surviving FDR correction. These findings support the oral–gut axis hypothesis and highlight the importance of maintaining natural dentition for gastrointestinal health.

## 1. Introduction

Tooth loss is a prevalent oral health condition among older adults, with substantial implications for nutritional intake, dietary patterns, and overall health^[1]^. The World Health Organization has identified oral health as an integral component of general health, and growing evidence suggests that poor oral health may be linked to systemic diseases, including cardiovascular disease, diabetes, and cognitive decline^[2; 3]^.

One potential pathway through which tooth loss may influence systemic health is via masticatory dysfunction. Reduced chewing efficiency has been associated with decreased intake of dietary fiber, fruits, and vegetables, and a shift toward softer, more processed foods^[4; 5]^. Such dietary changes may, in turn, alter gastrointestinal motility and bowel habits. Indeed, low fiber intake is a well-established risk factor for constipation, while dietary modifications may also contribute to diarrhea or irregular bowel patterns^[6]^. Furthermore, the oral cavity harbors a complex microbiome, and alterations in oral bacterial communities due to tooth loss may influence the gut microbiome through swallowed bacteria, potentially affecting intestinal function^[7; 8]^.

Despite these plausible biological mechanisms, epidemiological evidence directly linking objectively measured tooth loss to bowel habits remains limited. Large population-based surveys, including the China Health and Retirement Longitudinal Study (CHARLS) and the Health and Retirement Study (HRS), collect self-reported tooth loss data, which is susceptible to recall bias and lacks the precision of clinical examination. A recent study by Ji et al.^[9]^ used NHANES 2005–2008 data to examine the association between self-reported oral health and bowel habits assessed by the Bristol Stool Scale (BSS); however, that study did not utilize objective clinical dental examination data.

The NHANES 2005–2010 cycles are unique in that they simultaneously include a comprehensive clinical oral health examination (OHX) component and the Bristol Stool Scale assessment (BHQ060), providing an opportunity to examine the tooth loss–bowel habit relationship using objectively measured dental data. The present study aimed to investigate the association between the number of clinically examined missing teeth and abnormal bowel habits among US adults, using data from NHANES 2005–2010. We hypothesized that a greater number of missing teeth would be independently associated with an increased prevalence of abnormal bowel habits.

## 2. Materials and Methods

### 2.1 Study Design and Population

This cross-sectional study pooled data from three continuous NHANES cycles: 2005–2006, 2007–2008, and 2009–2010. NHANES is a nationally representative survey of the civilian, non-institutionalized US population, conducted by the National Center for Health Statistics (NCHS). The survey employs a complex, multistage probability sampling design and includes both interview and physical examination components^[10]^. The NHANES protocol was approved by the NCHS Research Ethics Review Board, and all participants provided written informed consent.

From an initial pool of 15,517 individuals with OHX and bowel health questionnaire records across the three cycles, we sequentially excluded: (1) 1,893 participants aged < 20 years; (2) 924 participants with missing or invalid BSS values (coded 77 or 99); (3) 370 pregnant women; (4) 1,222 participants with a history of cancer (MCQ220); and (5) 1,120 participants with missing covariate data. The final analytical sample comprised 9,988 adults. Missing data were handled using complete-case analysis, as the proportion of missing covariate data was low (approximately 10% of the eligible sample).

### 2.2 Exposure: Number of Missing Teeth

The primary exposure was the number of missing permanent teeth, assessed during the clinical oral health examination (OHX) conducted by trained dental examiners. Following the NHANES oral health examination protocol, each permanent tooth (excluding third molars, yielding a maximum of 28 teeth) was assessed, and tooth status was recorded. Teeth coded as 4 (missing due to caries) or 5 (missing due to other causes) were classified as missing^[11]^. Teeth coded as 3 (dental implant) were treated as non-missing in the primary analysis and excluded in a sensitivity analysis. Teeth coded as 9 (could not assess) were treated as non-missing. The total number of missing teeth was categorized into five groups: 0, 1–5, 6–10, 11–27, and 28 (complete edentulism).

### 2.3 Outcomes

#### 2.3.1 Bowel Habit Outcomes

Abnormal bowel habits was designated as the a priori primary outcome based on its clinical comprehensiveness and the validation of the BSS as a stool form assessment tool. Constipation and diarrhea were examined as secondary outcomes to decompose the primary finding. Three bowel habit outcomes were derived from the Bristol Stool Scale (BSS, variable BHQ060), a self-administered questionnaire component of the NHANES Bowel Health Questionnaire, which classifies stool form into seven types: Type 1 (separate hard lumps), Type 2 (sausage-shaped but lumpy), Type 3 (sausage with cracks on surface), Type 4 (smooth, soft sausage or snake), Type 5 (soft blobs with clear-cut edges), Type 6 (mushy, ragged edges), and Type 7 (watery, no solid pieces)^[12]^. Based on BSS, three bowel habit outcomes were defined:

1. Abnormal bowel habits (primary outcome): BSS 1–2 (constipation-type) or BSS 6–7 (diarrhea-type) versus BSS 3–5 (normal).
2. Constipation (secondary outcome): BSS 1–2 versus BSS 3–5.
3. Diarrhea (secondary outcome): BSS 6–7 versus BSS 3–5.

#### 2.3.2 Secondary Outcomes

Three secondary outcomes were derived from the Bowel Health Questionnaire, capturing components of fecal incontinence (bowel leakage). Participants who reported any accidental bowel leakage were asked about the composition of leakage. BHQ010, BHQ020, and BHQ040 assess bowel leakage of gas, mucus, and solid stool, respectively, using a 6-level frequency response scale: 1 = every day, 2 = every week, 3 = every month, 4 = a few times, 5 = once, and 6 = never. For the primary analysis of secondary outcomes, we used the “any” definition (responses 1–5 vs. 6) to maximize statistical power:

4. Gas leakage (BHQ010): Any occurrence of bowel leakage of gas (responses 1–5) versus never (response 6).
5. Mucus leakage (BHQ020): Any occurrence of bowel leakage of mucus (responses 1–5) versus never (response 6).
6. Solid stool leakage (BHQ040): Any occurrence of bowel leakage of solid stool (responses 1–5) versus never (response 6).

### 2.4 Covariates

Covariates were selected based on biological plausibility and prior literature to identify potential confounders in the tooth loss–bowel habit pathway. Demographic variables included age (continuous, RIDAGEYR), sex (male/female, RIAGENDR), race/ethnicity (non-Hispanic White, non-Hispanic Black, Mexican American, other [including Other Hispanic and Other Race]; RIDRETH1), education (≤high school, >high school; DMDEDUC2), and poverty-to-income ratio (PIR, continuous; INDFMPIR), a measure of family income relative to the federal poverty threshold. Health behavior variables included body mass index (BMI, continuous, measured during physical examination; BMXBMI), smoking status (never, former, current; derived from SMQ020 and SMQ040), alcohol consumption (drinker/non-drinker, defined as having had ≥12 drinks in any one year; ALQ101), and physical activity (active/inactive). Medical conditions included diabetes (yes/no; DIQ010), hypertension (yes/no; BPQ020), depression (PHQ-9 score ≥10, with PHQ-9 items DPQ010–DPQ090; for participants with 1–2 missing items, the total score was calculated from available items, and those with >2 missing items were excluded), and thyroid disease (yes/no; MCQ160M). Cancer history (MCQ220) was used as an exclusion criterion.

Physical activity was derived from NHANES physical activity questionnaires, which differed across cycles. For the 2007–2008 and 2009–2010 cycles, the Global Physical Activity Questionnaire (GPAQ) was used, with items PAQ605, PAQ620, PAQ635, PAQ650, and PAQ665 assessing moderate and vigorous physical activity across work, transportation, and recreational domains. For the 2005–2006 cycle, the GPAQ was not administered; instead, PAQ180 (self-rated daily activity level compared to peers) was used as a proxy, with responses of 1–2 (much more active / somewhat more active) classified as “active” and 3–5 (about the same / somewhat less / much less) classified as “inactive.” This approach ensured consistent active/inactive classification across all three cycles.

### 2.5 Statistical Analysis

All analyses incorporated NHANES survey design features, using the Mobile Examination Center (MEC) examination weights adjusted for three cycles (WTMEC2yr/3) to produce nationally representative estimates^[10]^. The survey design was specified with primary sampling units (PSU) defined by SDMVPSU, strata defined by SDMVSTRA, and nesting enabled. The option for lonely PSUs (strata with a single PSU) was set to “adjust” following CDC recommendations.

Hierarchical Testing Framework. We employed an a priori hierarchical testing strategy to address multiple comparisons without excessive loss of statistical power^[13; 14]^:

- Primary analysis (Tier 1): The primary outcome (abnormal bowel habits) was analyzed using P-for-trend across the five tooth loss categories (1 df test), using category-specific medians (0, 2, 8, 17, and 28 for groups 0, 1–5, 6–10, 11–27, and 28, respectively) as an ordinal variable. As the a priori primary hypothesis, this test was exempt from multiple comparison correction, consistent with established epidemiological practice holding that a priori primary hypotheses do not require adjustment[13; 14].
- Secondary analysis (Tier 2): Five secondary outcomes (constipation, diarrhea, gas leakage, mucus leakage, and solid stool leakage) were analyzed using P-for-trend. Benjamini– Hochberg FDR correction was applied across these 5 comparisons^[15]^, substantially reducing the multiplicity burden compared with the conventional approach of correcting across all pairwise category comparisons.
- Supplementary analysis (Tier 3): Exploratory analyses included: (a) continuous variable modeling (missing teeth as a continuous predictor, 1 df), estimating ORs per 1 and 5 additional missing teeth; (b) omnibus Wald tests for the joint significance of all 4 tooth loss category indicators (4 df), capturing the overall association including non-linear patterns; (c) restricted cubic splines (RCS) to model the dose–response relationship; and (d) a composite bowel outcome, defined as the presence of any abnormal bowel habit, gas leakage, mucus leakage, or solid stool leakage.

Regression Models. Survey-weighted logistic regression was used to estimate ORs and 95% CIs, with the quasibinomial family to accommodate potential overdispersion in the complex survey design. Confidence intervals were based on Wald-type statistics, and P-values were computed using design-based degrees of freedom (approximately 47 df, derived from the number of PSUs minus strata across the three pooled cycles). Three nested models were constructed: Model 1 (unadjusted); Model 2 (adjusted for age, sex, and race/ethnicity); and Model 3 (fully adjusted for age, sex, race/ethnicity, education, PIR, BMI, smoking, alcohol consumption, physical activity, diabetes, hypertension, depression, and thyroid disease). Multicollinearity was assessed using generalized variance inflation factors (GVIF), with all GVIF^(1/(2×Df)) values <1.4, well below the conventional threshold of 5, indicating no significant multicollinearity.

Dose–Response and Nonlinearity. Restricted cubic splines (RCS) with 4 degrees of freedom were employed to model the dose–response relationship between the number of missing teeth (continuous) and abnormal bowel habits, using natural splines (ns) with 3 interior knots at the 25th, 50th, and 75th percentiles. The overall significance of the spline association was assessed using a Wald test for the joint significance of all spline terms (4 df). The omnibus Wald test for the categorical model (4 df) provided a complementary assessment of the overall association. Subgroup Analyses. Subgroup analyses used a binary exposure (0–10 vs. 11–28 missing teeth), with the cutoff informed by the RCS-identified threshold and chosen at 10 to ensure adequate subgroup sample sizes. Subgroups included age (<60 vs. ≥60 years), sex, BMI (<30 vs. ≥30 kg/m²), diabetes status, and smoking status. Interactions were assessed using multiplicative interaction terms.

Sensitivity Analyses. Five sensitivity analyses were conducted: (1) excluding participants with dental implants (tooth status code 3); (2) redefining normal bowel habits as BSS 3–4 only; (3) redefining constipation as bowel movement frequency <3 times per week (BHD050); (4) adjusting for dietary fiber intake (g/day, from NHANES 24-hour dietary recall) as an additional covariate; and (5) adjusting for denture use (removable partial or full denture, from OHQ410 and OHQ430) among participants from NHANES 2005–2008 cycles, where denture use data were available.

All statistical analyses were performed using R software version 4.5.2 (R Foundation for Statistical Computing, Vienna, Austria), with the survey package^[16]^ for survey-weighted analyses and the splines package for RCS modeling. A two-sided P value <0.05 was considered statistically significant. This study was reported following the STROBE guidelines for cross-sectional studies.

## 3. Results

### 3.1 Study Population

The participant selection process is illustrated in Figure 1. From the initial 15,517 individuals with OHX and bowel health records across three NHANES cycles (2005–2010), 1,893 were excluded for being aged < 20 years, 924 for missing or invalid BSS values, 370 for pregnancy, 1,222 for cancer history, and 1,120 for missing covariates. The final analytical sample comprised 9,988 adults.

**Figure 1.**
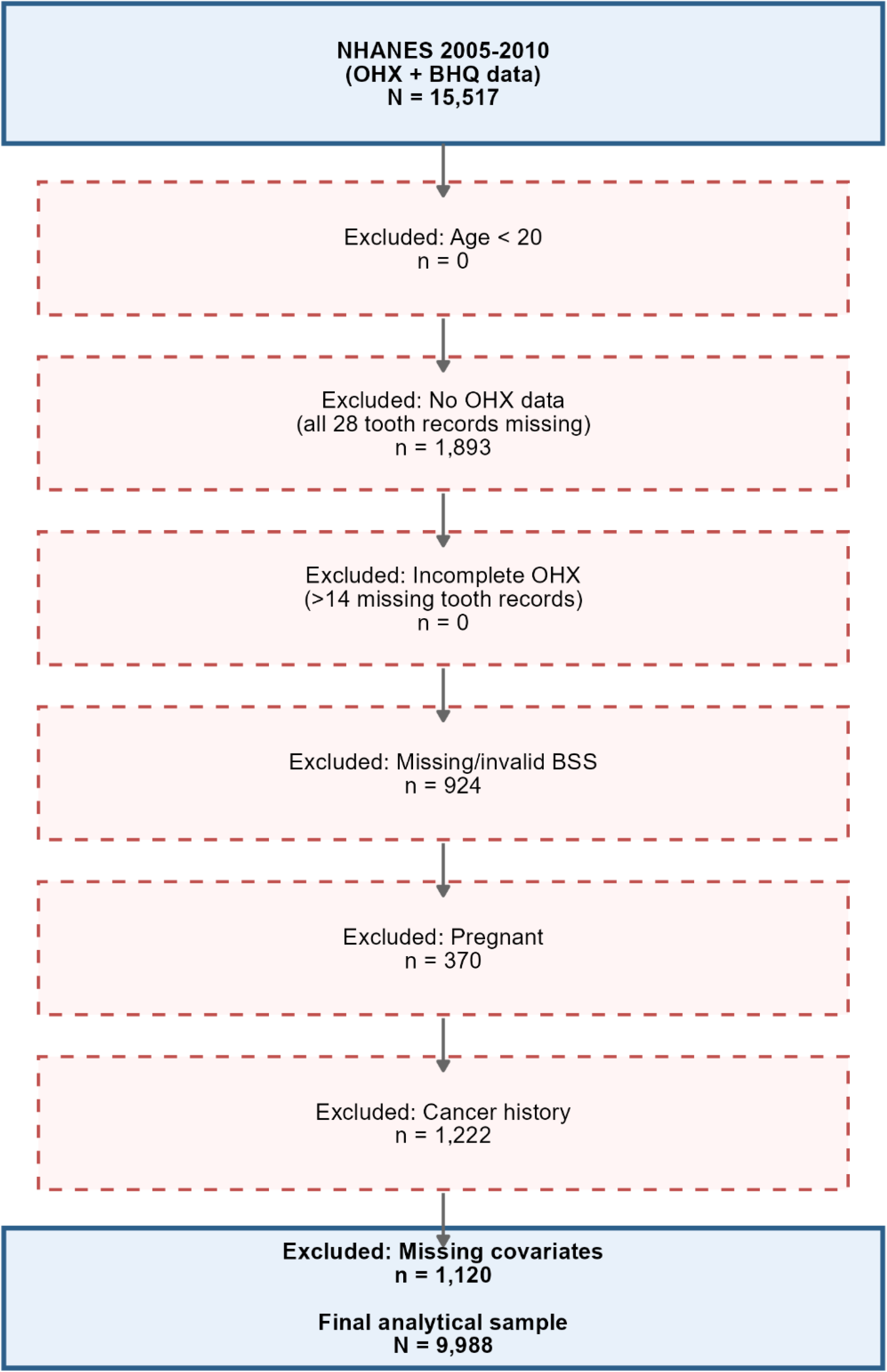
Flow chart of participant selection from NHANES 2005–2010.

**Figure 2.**
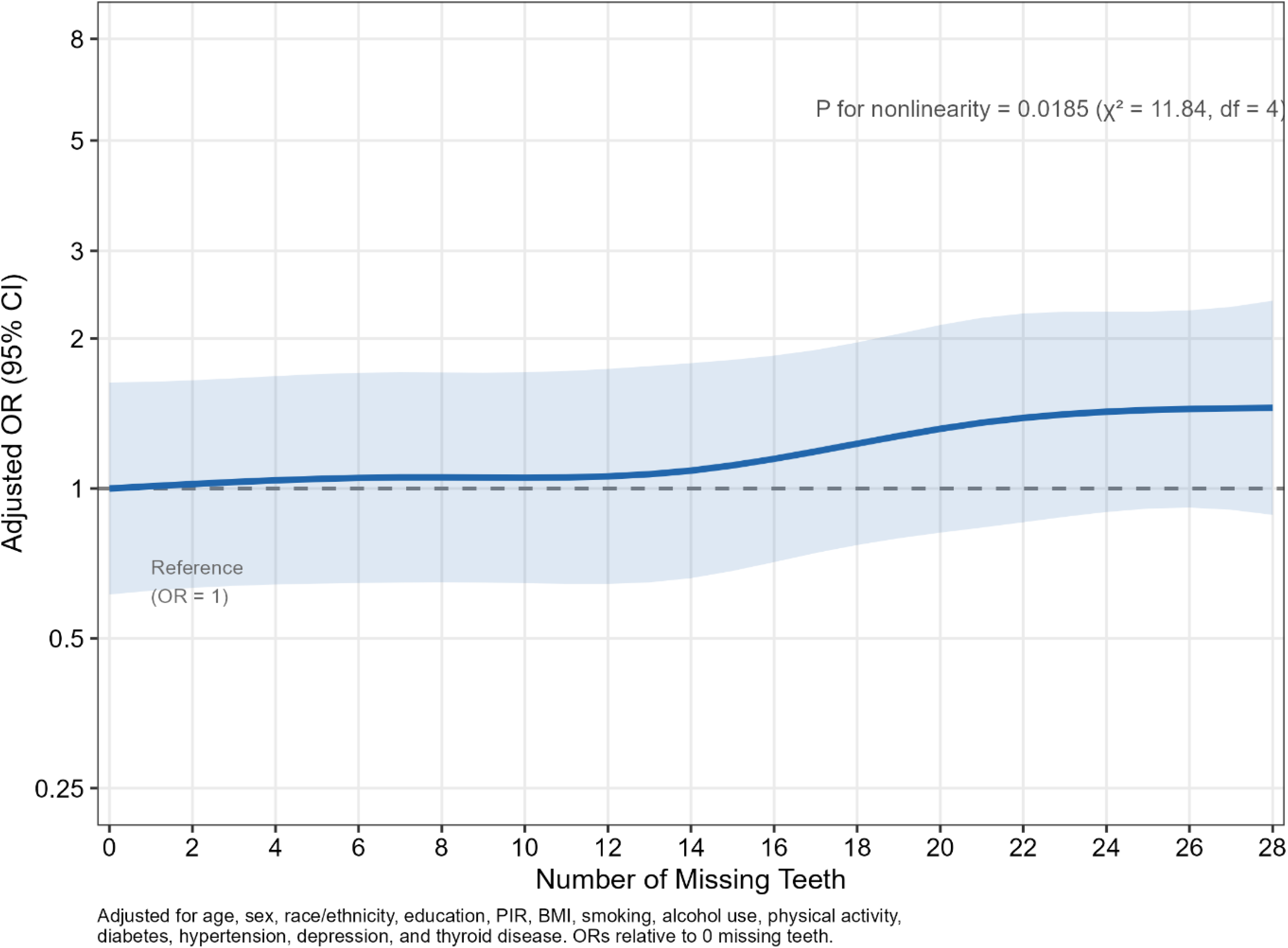
Restricted cubic spline analysis of the association between number of missing teeth (continuous) and abnormal bowel habits. The solid line represents the adjusted odds ratio, and the shaded area represents the 95% confidence interval. The reference value is set at 0 missing teeth. Overall spline test: χ² = 11.84, df = 4, P = 0.019. Omnibus Wald test for categorical model: χ² = 14.40, df = 4, P = 0.006.

**Figure 3.**
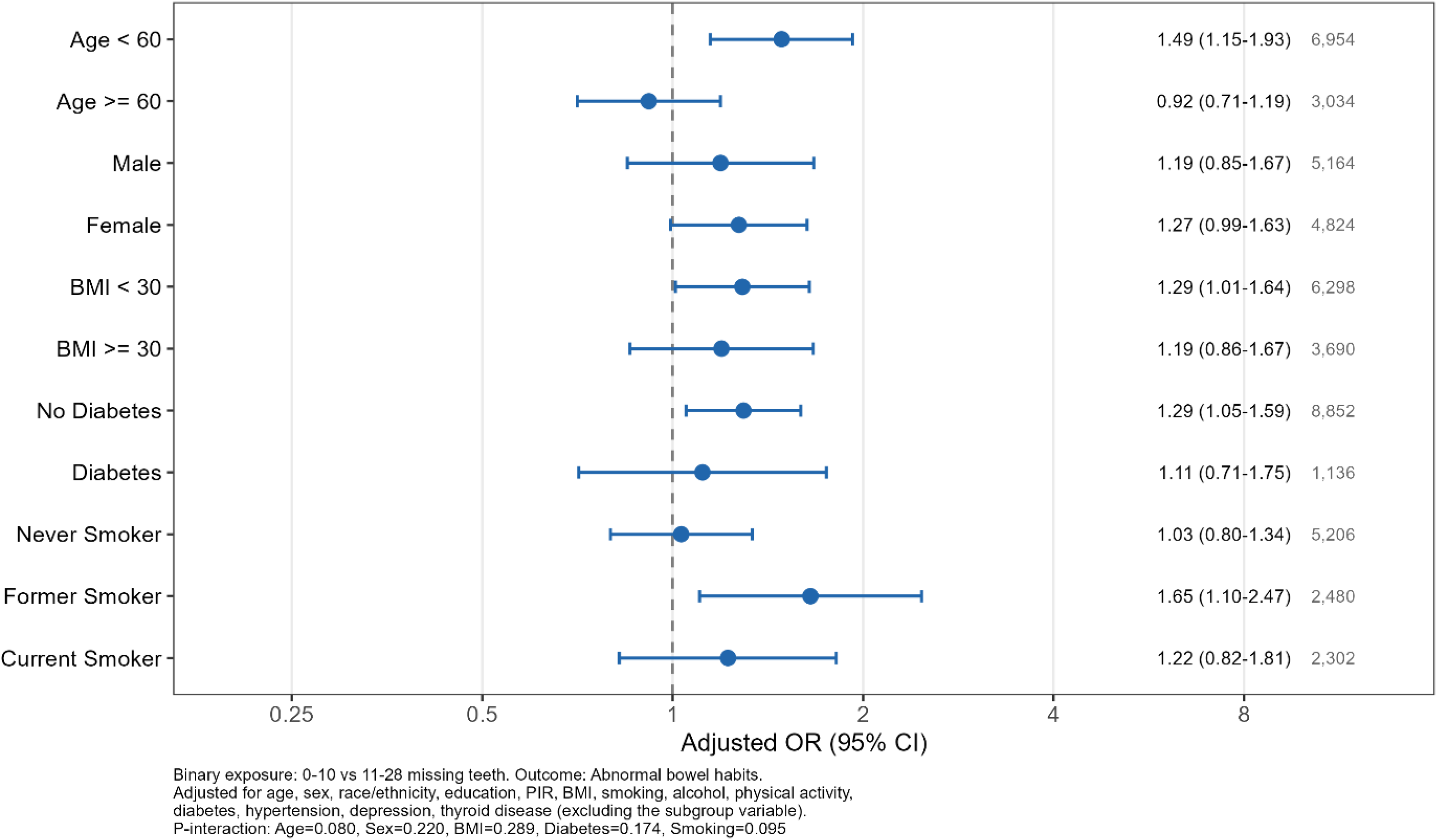
Forest plot of subgroup analyses for the association between tooth loss (11–28 vs. 0–10 missing teeth) and abnormal bowel habits. Subgroups include age (<60 vs. ≥60 years), sex, BMI (<30 vs. ≥30 kg/m²), diabetes status, and smoking status (current, former, never). Squares represent odds ratios with horizontal lines indicating 95% confidence intervals. P-interaction values are shown for each subgroup. All models were fully adjusted for age, sex, race/ethnicity, education, PIR, BMI, smoking, alcohol consumption, physical activity, diabetes, hypertension, depression, and thyroid disease (with the subgroup variable removed from covariates where applicable).

### 3.2 Baseline Characteristics

The distribution of missing teeth was as follows: 0 teeth missing (n = 3,334), 1–5 (n = 3,450), 6– 10 (n = 1,168), 11–27 (n = 1,283), and 28 (n = 753). Table 1 presents the baseline characteristics of the study population stratified by tooth loss category.

**Table 1.** Baseline Characteristics of Study Population by Number of Missing Teeth, NHANES 2005–2010 (N = 9,988)

| Variable | Teeth = 0<br>(n = 3,334) | Teeth = 1–5<br>(n = 3,450) | Teeth = 6–<br>10 (n =<br>1,168) | Teeth = 11–<br>27 (n =<br>1,283) | Teeth = 28<br>(n = 753) | P<br>value |
| --- | --- | --- | --- | --- | --- | --- |
| **Continuous variables**<br>**(weighted mean ± SE)** |  |  |  |  |  |  |
| Age (years) | 39.2 ± 0.3 | 46.8 ± 0.4 | 54.0 ± 0.6 | 60.5 ± 0.4 | 63.6 ± 0.7 | <0.001 |
| PIR | 3.4 ± 0.1 | 3.2 ± 0.0 | 2.7 ± 0.1 | 2.3 ± 0.1 | 2.3 ± 0.1 | <0.001 |
| BMI (kg/m<sup>2</sup>) | 28.0 ± 0.2 | 28.9 ± 0.2 | 29.9 ± 0.3 | 29.2 ± 0.2 | 29.2 ± 0.3 | <0.001 |
| Missing teeth (n) | 0.0 ± 0.0 | 2.6 ± 0.0 | 7.6 ± 0.1 | 17.3 ± 0.2 | 28.0 ± 0.0 | <0.001 |
| **Categorical variables**<br>**(weighted %)** |  |  |  |  |  |  |
| Male | 54.0 | 48.0 | 47.4 | 50.8 | 43.4 | <0.001 |
| NH White | 75.7 | 69.3 | 63.0 | 67.0 | 78.6 | <0.001 |
| NH Black | 7.7 | 10.9 | 17.5 | 19.3 | 12.0 | <0.001 |
| Mexican American | 8.2 | 9.2 | 7.4 | 5.6 | 2.8 | <0.001 |
| Other race | 8.4 | 10.6 | 12.1 | 8.2 | 6.6 | <0.001 |
| >High school education | 72.3 | 56.3 | 43.4 | 31.0 | 25.0 | <0.001 |
| Never smoker | 61.4 | 53.1 | 44.6 | 35.9 | 27.4 | <0.001 |
| Former smoker | 20.9 | 24.7 | 25.0 | 30.0 | 36.7 | <0.001 |
| Current smoker | 17.7 | 22.2 | 30.4 | 34.0 | 35.9 | <0.001 |
| Drinker | 82.0 | 76.1 | 71.2 | 66.8 | 65.3 | <0.001 |
| Physical activity (Active) | 79.9 | 75.0 | 70.1 | 70.5 | 67.8 | <0.001 |
| Diabetes | 3.6 | 7.4 | 12.5 | 16.9 | 22.5 | <0.001 |
| Hypertension | 18.0 | 30.3 | 43.4 | 48.4 | 54.0 | <0.001 |
| Depression (PHQ-9 ≥ 10) | 4.4 | 6.6 | 9.5 | 9.9 | 11.7 | <0.001 |
| Thyroid disease | 6.1 | 10.4 | 13.3 | 12.3 | 16.5 | <0.001 |

*Note: PIR = poverty-to-income ratio; BMI = body mass index; PHQ-9 = Patient Health*
*Questionnaire-9. Continuous variables compared using survey-weighted ANOVA; categorical variables compared using survey-weighted chi-square tests.*

Participants with more missing teeth were significantly older (mean age: 39.2 ± 0.3 years for those with 0 missing teeth vs. 63.6 ± 0.7 years for those with 28 missing teeth; P < 0.001), had lower PIR (3.4 ± 0.1 vs. 2.3 ± 0.1; P < 0.001), and had higher BMI (28.0 ± 0.2 vs. 29.2 ± 0.3 kg/m²; P < 0.001). The mean number of missing teeth ranged from 0.0 ± 0.0 in the reference group to 28.0 ± 0.0 in the edentulous group.

Regarding categorical variables, participants with complete tooth loss (28 missing teeth) compared to those with no missing teeth showed lower proportions of males (43.4% vs. 54.0%; P < 0.001), higher proportions of non-Hispanic Black individuals (12.0% vs. 7.7%; P < 0.001), substantially lower educational attainment (>high school: 25.0% vs. 72.3%; P < 0.001), higher smoking rates (current smoker: 35.9% vs. 17.7%; former smoker: 36.7% vs. 20.9%; P < 0.001), and higher prevalence of diabetes (22.5% vs. 3.6%; P < 0.001), hypertension (54.0% vs. 18.0%; P < 0.001), depression (PHQ-9 ≥10: 11.7% vs. 4.4%; P < 0.001), and thyroid disease (16.5% vs. 6.1%; P < 0.001). Physical activity was lower among edentulous participants (active: 67.8% vs. 79.9%; P < 0.001), and non-drinking was more common (34.7% vs. 18.0%; P < 0.001).

### 3.3 Outcome Distribution

The BSS distribution was: Type 1 (n = 201), Type 2 (n = 508), Type 3 (n = 2,419), Type 4 (n = 5,314), Type 5 (n = 804), Type 6 (n = 652), and Type 7 (n = 90). The prevalence of primary outcomes was: abnormal bowel habits 1,451 (14.5%), constipation 709 (7.1%), and diarrhea 742 (7.4%). For secondary outcomes, using the “any” definition (responses 1–5 vs. 6), the prevalence was: any gas leakage 4,334 (43.5%), any mucus leakage 311 (3.1%), and any solid stool leakage 197 (2.0%).

### 3.4 Association Between Missing Teeth and Outcomes

Table 2 presents the categorical survey-weighted logistic regression results for all six outcomes across three models. The primary analysis (P-for-trend) and supplementary analyses (continuous variable, omnibus Wald test, RCS) are summarized in Table 5.

**Table 2.**
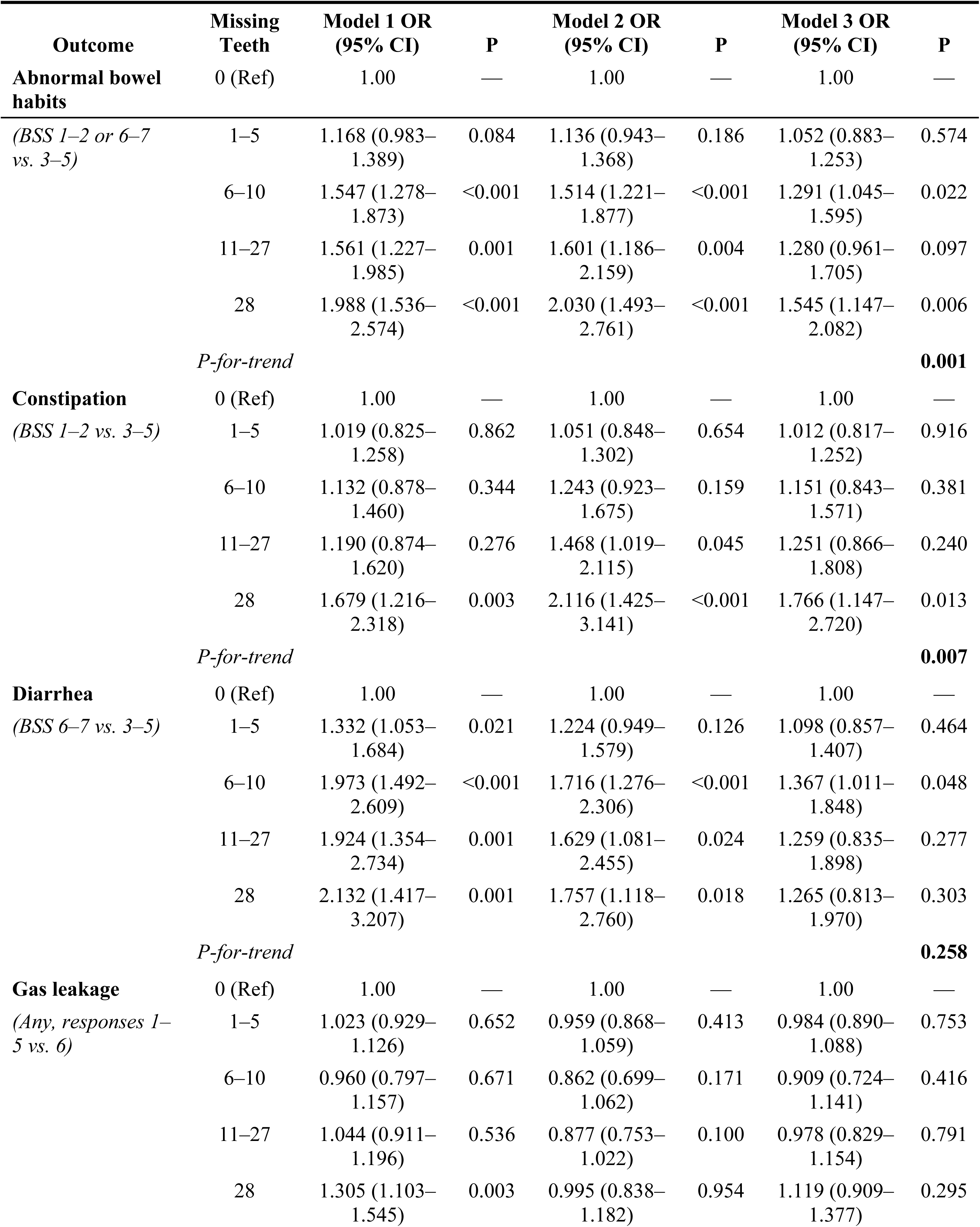
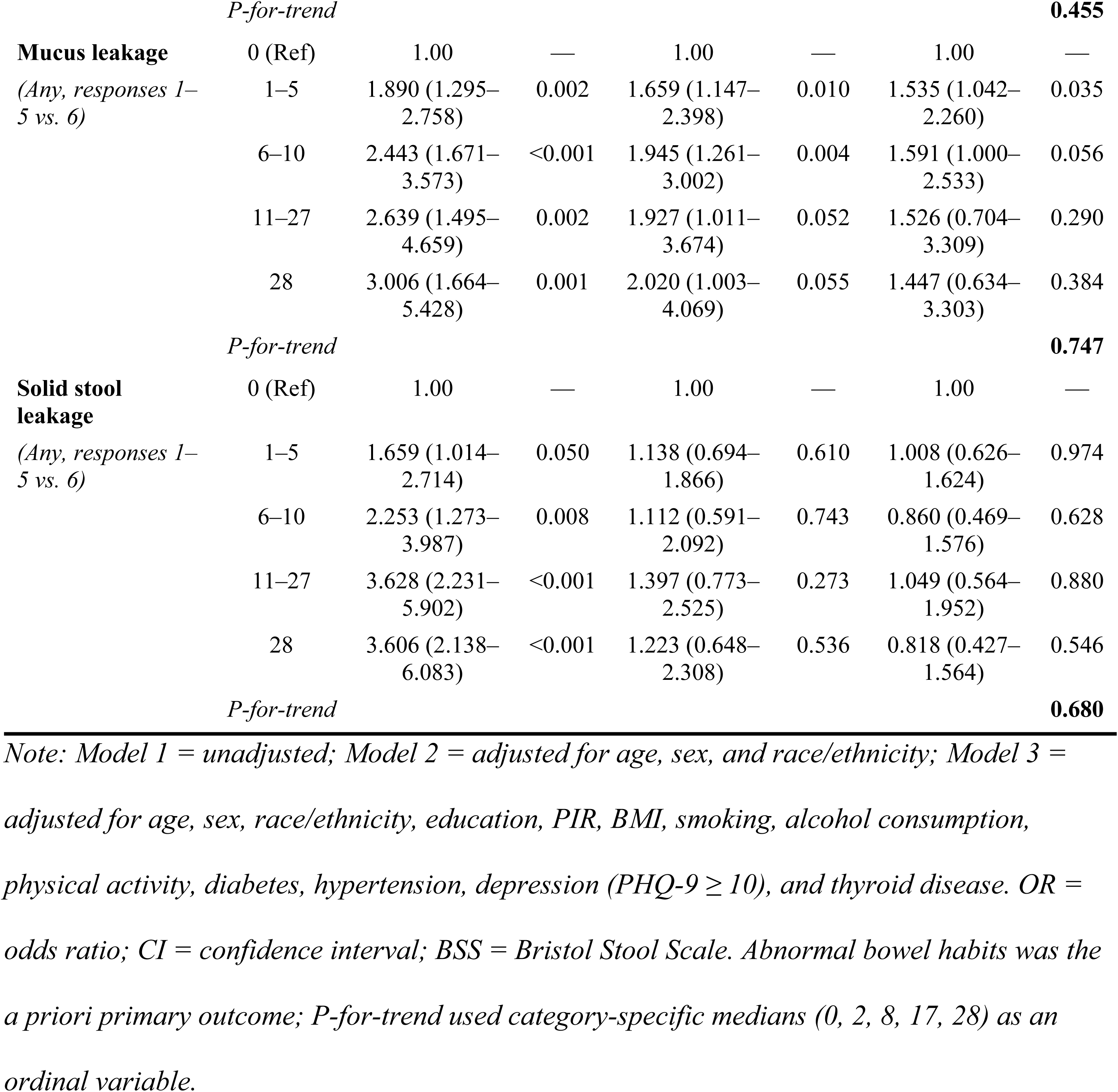
Survey-Weighted Logistic Regression for the Association Between Number of Missing Teeth and Bowel Habit Outcomes, NHANES 2005–2010.

**Table 3.** Subgroup Analysis: Association Between Tooth Loss (11–28 vs. 0–10 Missing Teeth) and Abnormal Bowel Habits.

| Subgroup | OR (95% CI) | P value | P-interaction |
| --- | --- | --- | --- |
| **Age** |  |  | 0.080 |
| <60 years | 1.486 (1.147–1.926) | 0.004 |  |
| ≥60 years | 0.917 (0.706–1.190) | 0.516 |  |
| **Sex** |  |  | 0.220 |
| Female | 1.273 (0.993–1.631) | 0.063 |  |
| Male | 1.190 (0.848–1.671) | 0.320 |  |
| **BMI** |  |  | 0.289 |
| <30 kg/m<sup>2</sup> | 1.289 (1.010–1.644) | 0.047 |  |
| ≥30 kg/m<sup>2</sup> | 1.194 (0.855–1.668) | 0.302 |  |
| **Diabetes** |  |  | 0.174 |
| No | 1.294 (1.051–1.594) | 0.019 |  |
| Yes | 1.115 (0.710–1.749) | 0.639 |  |
| **Smoking** |  |  | 0.095 |
| Current smoker | 1.223 (0.824–1.814) | 0.323 |  |
| Former smoker | 1.651 (1.102–2.474) | 0.019 |  |
| Never smoker | 1.032 (0.797–1.336) | 0.813 |  |

*Note: Analyses used survey-weighted logistic regression with binary exposure (0–10 vs. 11–28 missing teeth), fully adjusted for all covariates. P-interaction assessed using multiplicative interaction terms. BMI = body mass index.*

**Table 4.**
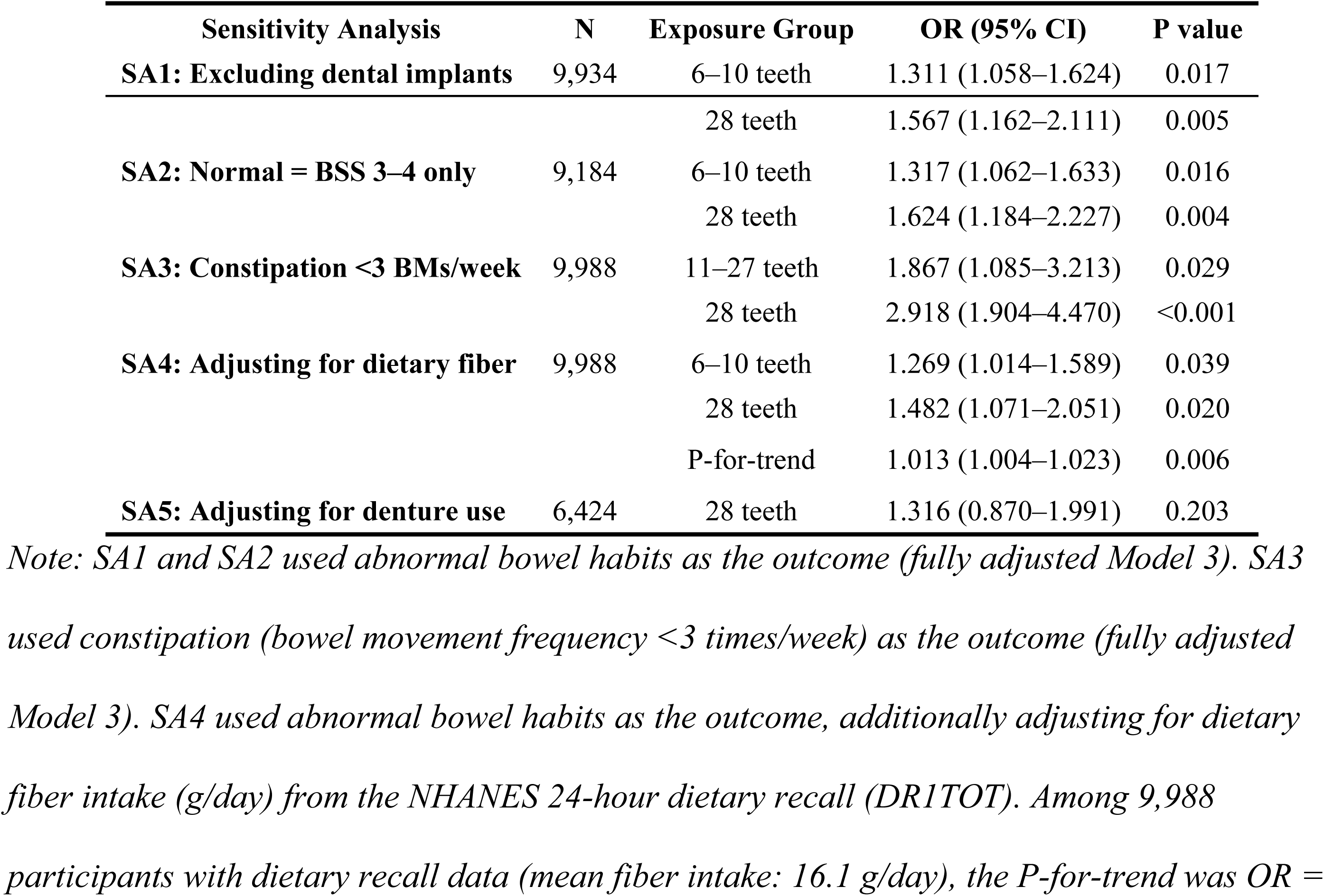
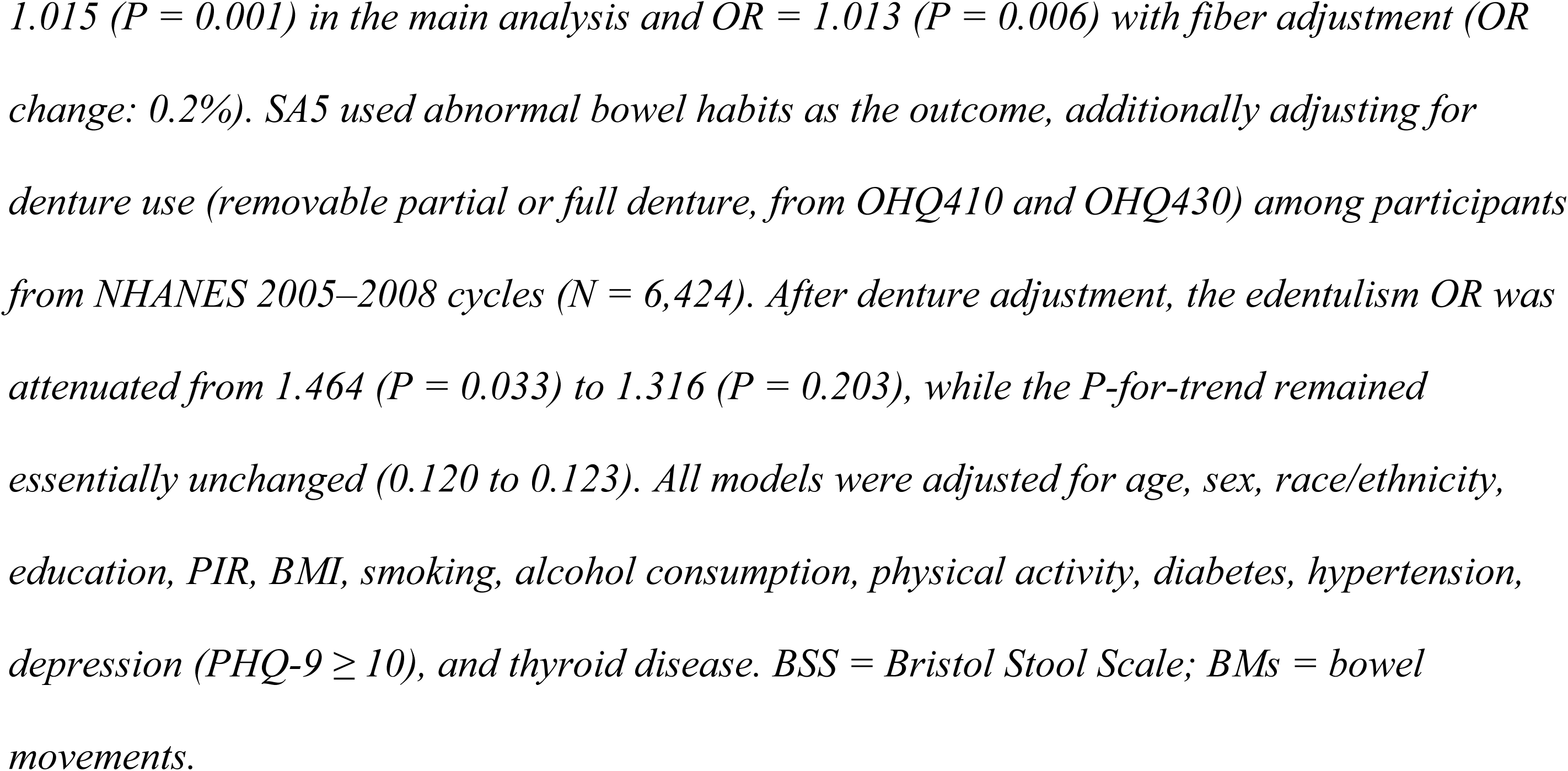
Sensitivity Analyses for the Association Between Tooth Loss and Primary Outcomes.

| Sensitivity Analysis | N | Exposure Group | OR (95% CI) | P value |
| --- | --- | --- | --- | --- |
| **SA1: Excluding dental implants** | 9,934 | 6–10 teeth | 1.311 (1.058–1.624) | 0.017 |
|  |  | 28 teeth | 1.567 (1.162–2.111) | 0.005 |
| **SA2: Normal = BSS 3–4 only** | 9,184 | 6–10 teeth | 1.317 (1.062–1.633) | 0.016 |
|  |  | 28 teeth | 1.624 (1.184–2.227) | 0.004 |
| **SA3: Constipation <3 BMs/week** | 9,988 | 11–27 teeth | 1.867 (1.085–3.213) | 0.029 |
|  |  | 28 teeth | 2.918 (1.904–4.470) | <0.001 |
| **SA4: Adjusting for dietary fiber** | 9,988 | 6–10 teeth | 1.269 (1.014–1.589) | 0.039 |
|  |  | 28 teeth | 1.482 (1.071–2.051) | 0.020 |
|  |  | P-for-trend | 1.013 (1.004–1.023) | 0.006 |
| **SA5: Adjusting for denture use** | 6,424 | 28 teeth | 1.316 (0.870–1.991) | 0.203 |

*Note: SA1 and SA2 used abnormal bowel habits as the outcome (fully adjusted Model 3). SA3 used constipation (bowel movement frequency <3 times/week) as the outcome (fully adjusted Model 3). SA4 used abnormal bowel habits as the outcome, additionally adjusting for dietary fiber intake (g/day) from the NHANES 24-hour dietary recall (DRITOT). Among 9,988 participants with dietary recall data (mean fiber intake: 16.1 g/day), the P-for-trend was OR =*
1.015 ( $P = 0.001$ ) in the main analysis and $OR = 1.013$ ( $P = 0.006$ ) with fiber adjustment ( $OR$ change: 0.2%). SA5 used abnormal bowel habits as the outcome, additionally adjusting for denture use (removable partial or full denture, from OHQ410 and OHQ430) among participants from NHANES 2005–2008 cycles ( $N = 6,424$ ). After denture adjustment, the edentulism $OR$ was attenuated from 1.464 ( $P = 0.033$ ) to 1.316 ( $P = 0.203$ ), while the $P$ -for-trend remained essentially unchanged (0.120 to 0.123). All models were adjusted for age, sex, race/ethnicity, education, PIR, BMI, smoking, alcohol consumption, physical activity, diabetes, hypertension, depression ( $PHQ-9 \geq 10$ ), and thyroid disease. BSS = Bristol Stool Scale; BMs = bowel movements.

**Table 5.**
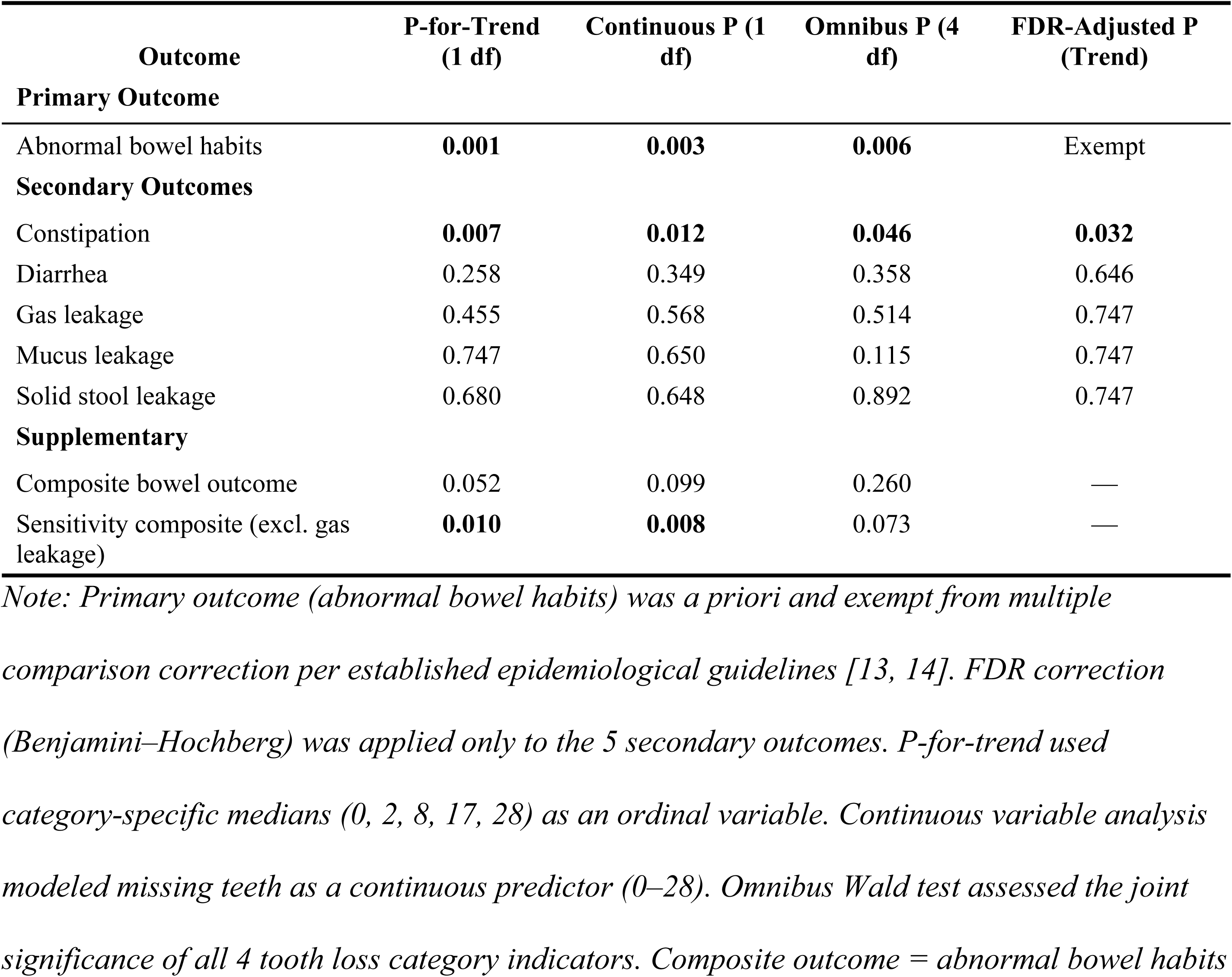
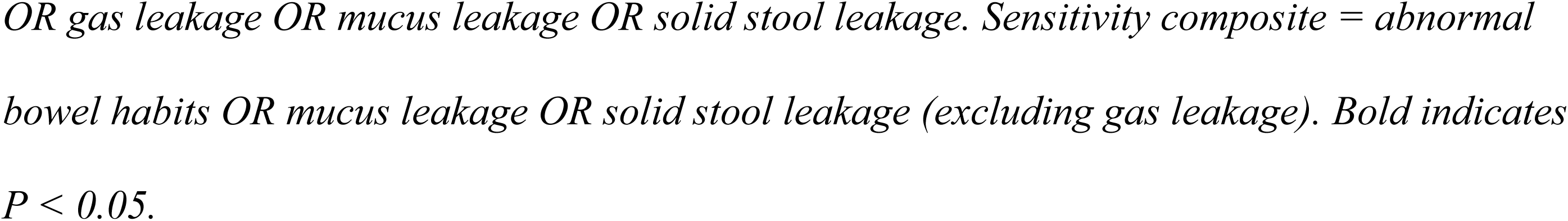
Summary of Hierarchical Testing Framework Results, NHANES 2005–2010.

| Outcome | P-for-Trend<br>(1 df) | Continuous P (1<br>df) | Omnibus P (4<br>df) | FDR-Adjusted P<br>(Trend) |
| --- | --- | --- | --- | --- |
| **Primary Outcome** |  |  |  |  |
| Abnormal bowel habits | **0.001** | **0.003** | **0.006** | Exempt |
| **Secondary Outcomes** |  |  |  |  |
| Constipation | **0.007** | **0.012** | **0.046** | **0.032** |
| Diarrhea | 0.258 | 0.349 | 0.358 | 0.646 |
| Gas leakage | 0.455 | 0.568 | 0.514 | 0.747 |
| Mucus leakage | 0.747 | 0.650 | 0.115 | 0.747 |
| Solid stool leakage | 0.680 | 0.648 | 0.892 | 0.747 |
| **Supplementary** |  |  |  |  |
| Composite bowel outcome | 0.052 | 0.099 | 0.260 | — |
| Sensitivity composite (excl. gas leakage) | **0.010** | **0.008** | 0.073 | — |

*Note: Primary outcome (abnormal bowel habits) was a priori and exempt from multiple comparison correction per established epidemiological guidelines [13, 14]. FDR correction (Benjamini–Hochberg) was applied only to the 5 secondary outcomes. P-for-trend used category-specific medians (0, 2, 8, 17, 28) as an ordinal variable. Continuous variable analysis modeled missing teeth as a continuous predictor (0–28). Omnibus Wald test assessed the joint significance of all 4 tooth loss category indicators. Composite outcome = abnormal bowel habits*
*OR gas leakage OR mucus leakage OR solid stool leakage. Sensitivity composite = abnormal bowel habits OR mucus leakage OR solid stool leakage (excluding gas leakage). Bold indicates $P < 0.05$ .*

#### 3.4.1 Categorical Analysis

In fully adjusted models (Model 3), both the 6–10 missing teeth group (OR = 1.291, 95% CI: 1.045–1.595, P = 0.022) and complete edentulism (28 missing teeth; OR = 1.545, 95% CI: 1.147–2.082, P = 0.006) were significantly associated with abnormal bowel habits. For constipation, complete edentulism was significantly associated (OR = 1.766, 95% CI: 1.147– 2.720, P = 0.013). For diarrhea, the 6–10 missing teeth group showed a significant association (OR = 1.367, 95% CI: 1.011–1.848, P = 0.048). Among secondary outcomes, mucus leakage showed a significant association for the 1–5 missing teeth group (OR = 1.535, 95% CI: 1.042– 2.260, P = 0.035), while gas leakage and solid stool leakage showed no significant associations in fully adjusted models.

#### 3.4.2 Primary Analysis: P-for-Trend

In the a priori primary analysis, the P-for-trend across tooth loss categories was significant for abnormal bowel habits (OR per category median = 1.015, 95% CI: 1.006–1.024, P = 0.001). For constipation, the trend was also significant (OR = 1.019, 95% CI: 1.006–1.033, P = 0.007). No significant trends were observed for diarrhea (P = 0.258), gas leakage (P = 0.455), mucus leakage (P = 0.747), or solid stool leakage (P = 0.680).

#### 3.4.3 Supplementary: Continuous Variable Analysis

When missing teeth was modeled as a continuous variable, each additional missing tooth was associated with a 1.4% increase in the odds of abnormal bowel habits (OR = 1.014, 95% CI: 1.005–1.023, P = 0.003). For clinical interpretation, each 5 additional missing teeth was associated with OR = 1.071 (95% CI: 1.027–1.117). For constipation, each additional missing tooth was associated with a 1.9% increase in odds (OR = 1.019, 95% CI: 1.005–1.033, P = 0.012; OR per 5 teeth = 1.099, 95% CI: 1.024–1.179). No significant continuous associations were observed for diarrhea (P = 0.349), gas leakage (P = 0.568), mucus leakage (P = 0.650), or solid stool leakage (P = 0.648).

#### 3.4.4 Supplementary: Omnibus Wald Test

The omnibus Wald test, assessing the overall association across all 4 tooth loss category indicators (4 df), was significant for abnormal bowel habits (χ² = 14.40, df = 4, P = 0.006) and for constipation (χ² = 9.70, df = 4, P = 0.046). The omnibus test for mucus leakage was non-significant (χ² = 7.42, df = 4, P = 0.115), while diarrhea (P = 0.358), gas leakage (P = 0.514), and solid stool leakage (P = 0.892) were non-significant.

### 3.5 Secondary Outcomes and FDR Correction

Secondary outcomes (constipation, diarrhea, gas leakage, mucus leakage, and solid stool leakage) were analyzed using P-for-trend, with Benjamini–Hochberg FDR correction applied across 5 comparisons. The categorical results are presented in Table 2.

After FDR correction, constipation reached statistical significance (FDR-adjusted P = 0.032), while diarrhea (FDR-adjusted P = 0.646), gas leakage (FDR-adjusted P = 0.747), mucus leakage (FDR-adjusted P = 0.747), and solid stool leakage (FDR-adjusted P = 0.747) did not reach significance. The survival of constipation after FDR correction provides specific evidence for the tooth loss–constipation pathway and supports the biological plausibility of the masticatory dysfunction hypothesis. The remaining secondary outcomes are considered exploratory and should be interpreted with caution.

### 3.6 Composite Bowel Outcome

A composite bowel outcome was defined as the presence of any of: abnormal bowel habits, any gas leakage (responses 1–5), any mucus leakage (responses 1–5), or any solid stool leakage (responses 1–5). The “any” definition was used for all components to maximize sensitivity.

Among 9,988 participants, 5,126 (51.3%) had at least one bowel symptom. The high prevalence of the composite outcome reflects the inclusion of the broad “any” gas leakage definition (43.5% prevalence); abnormal bowel habits, mucus leakage, and solid stool leakage contributed 14.5%, 3.1%, and 2.0%, respectively.

In fully adjusted categorical models, complete edentulism was significantly associated with the composite outcome (OR = 1.269, 95% CI: 1.017–1.583, P = 0.041). The P-for-trend was of borderline significance (OR per category median = 1.008, 95% CI: 1.000–1.015, P = 0.052), as was the continuous variable analysis (OR per 1 tooth = 1.006, P = 0.099). The omnibus Wald test was not significant (χ² = 5.28, df = 4, P = 0.260). The borderline results for the composite including gas leakage reflect the dominance of the high-prevalence gas leakage component, which was not associated with tooth loss.

Because the broad “any” gas leakage definition (43.5% prevalence) dominated the composite outcome, a sensitivity composite excluding gas leakage was also examined. This sensitivity composite (abnormal bowel habits, mucus leakage, or solid stool leakage) yielded a prevalence of 17.8% (n = 1,781) and showed significant associations with tooth loss: in categorical models, participants with 6–10 missing teeth (OR = 1.277, 95% CI: 1.018–1.602, P = 0.040) and those with complete edentulism (OR = 1.425, 95% CI: 1.065–1.908, P = 0.021) had significantly higher odds. The P-for-trend was significant (OR per category median = 1.013, 95% CI: 1.003– 1.022, P = 0.010), as was the continuous variable analysis (OR per 1 tooth = 1.012, 95% CI: 1.004–1.021, P = 0.008; OR per 5 teeth = 1.062, 95% CI: 1.018–1.109). The omnibus Wald test was of borderline significance (χ² = 8.57, df = 4, P = 0.073).

### 3.7 Dose–Response Relationship: Restricted Cubic Splines

Restricted cubic splines with 4 degrees of freedom were used to model the relationship between the number of missing teeth (continuous) and abnormal bowel habits. The Wald test for the overall significance of the spline association yielded χ² = 11.84 (df = 4, P = 0.019), indicating a statistically significant nonlinear dose–response relationship. The spline curve showed that the odds ratio remained near 1.0 from 0 to approximately 10 missing teeth, after which it began to rise gradually. At 28 missing teeth, the predicted OR was 1.452 (95% CI: 0.885–2.382). The significant spline test, together with the significant omnibus Wald test for the categorical model (χ² = 14.40, df = 4, P = 0.006), provides strong evidence for a dose–response relationship between tooth loss and abnormal bowel habits.

### 3.8 Subgroup Analysis

Subgroup analyses were performed using a binary exposure (0–10 vs. 11–28 missing teeth) with abnormal bowel habits as the outcome (Table 3). Among participants aged <60 years, those with 11–28 missing teeth had significantly higher odds of abnormal bowel habits (OR = 1.486, 95% CI: 1.147–1.926, P = 0.004), whereas no association was observed among those aged ≥60 years (OR = 0.917, 95% CI: 0.706–1.190, P = 0.516; P-interaction = 0.080).

Among former smokers, the association between tooth loss and abnormal bowel habits was significant (OR = 1.651, 95% CI: 1.102–2.474, P = 0.019), while current smokers (OR = 1.223, 95% CI: 0.824–1.814, P = 0.323) and never smokers (OR = 1.032, 95% CI: 0.797–1.336, P = 0.813) showed no significant associations (P-interaction = 0.095).

Among non-diabetic participants, the association was significant (OR = 1.294, 95% CI: 1.051– 1.594, P = 0.019), while diabetic participants showed no significant association (OR = 1.115, 95% CI: 0.710–1.749, P = 0.639; P-interaction = 0.174). No significant interactions were observed by sex (P-interaction = 0.220) or BMI (P-interaction = 0.289). Although elevated BMI has been shown to alter colonic sensory-motor function and transit^[17]^, the similar associations across BMI strata suggest that obesity-related physiological changes do not substantially modify the tooth loss–bowel habit relationship.

### 3.9 Sensitivity Analyses

Five sensitivity analyses were conducted to assess the robustness of the primary finding (Table 4), the fourth and fifth of which (dietary fiber and denture use adjustments) are discussed in detail in Section 4.6:

1. Excluding participants with dental implants (N = 9,934): The associations for both 6–10 missing teeth (OR = 1.311, 95% CI: 1.058–1.624, P = 0.017) and complete edentulism (OR = 1.567, 95% CI: 1.162–2.111, P = 0.005) remained significant.
2. Redefining normal bowel habits as BSS 3–4 only (N = 9,184): The associations for 6–10 missing teeth (OR = 1.317, 95% CI: 1.062–1.633, P = 0.016) and complete edentulism (OR = 1.624, 95% CI: 1.184–2.227, P = 0.004) remained significant.
3. Redefining constipation as bowel movement frequency <3 times/week (N = 9,988, prevalence 3.3%): The associations for 11–27 missing teeth (OR = 1.867, 95% CI: 1.085–3.213, P = 0.029) and complete edentulism (OR = 2.918, 95% CI: 1.904–4.470, P < 0.001) were highly significant.
4. Adjusting for dietary fiber intake (N = 9,988): After additional adjustment for dietary fiber (g/day), the P-for-trend remained significant (OR = 1.013, 95% CI: 1.004–1.023, P = 0.006), with minimal attenuation of individual category ORs (e.g., 28 teeth: OR = 1.545 → 1.482). Dietary fiber itself was not significantly associated with abnormal bowel habits (P = 0.089).
5. Adjusting for denture use (NHANES 2005–2008 subset, N = 6,424): Denture use data (OHQ410 and OHQ430) were available only for NHANES 2005–2008 cycles. Among the 6,424 participants with denture data, 1,458 (22.7%) reported using a removable partial or full denture, with prevalence strongly correlated with tooth loss severity (0.3% among those with 0 missing teeth vs. 93.3% among edentulous participants). In this two-cycle subset, the unadjusted association (Model A) showed an OR of 1.464 (95% CI: 1.047–2.048, P = 0.033) for edentulism and a P-for-trend of 0.120. After additional adjustment for denture use (Model B), the edentulism OR was modestly attenuated to 1.316 (95% CI: 0.870–1.991, P = 0.203), with minimal change in P-for-trend (0.123). Denture use itself was not independently associated with abnormal bowel habits (OR = 1.124, 95% CI: 0.859–1.471, P = 0.401). The approximately 10% attenuation of the edentulism effect, combined with the unchanged P-for-trend and the non-significant independent effect of denture use, suggests that denture use partially confounds but does not fully explain the observed association.

## 4. Discussion

### 4.1 Principal Findings

This cross-sectional study using NHANES 2005–2010 data employed an a priori hierarchical testing framework to examine the association between clinically measured tooth loss and abnormal bowel habits. In the primary analysis, the P-for-trend across tooth loss categories was significant (OR per category median = 1.015, 95% CI: 1.006–1.024, P = 0.001), indicating a dose–response relationship between increasing tooth loss and abnormal bowel habits. This finding was corroborated by multiple complementary analyses: the omnibus Wald test (P = 0.006), continuous variable analysis (P = 0.003), and restricted cubic spline analysis (P = 0.019). A sensitivity composite bowel outcome excluding gas leakage also showed significant associations (P-for-trend = 0.010, continuous P = 0.008). In categorical models, both the 6–10 missing teeth group (OR = 1.291, P = 0.022) and complete edentulism (OR = 1.545, P = 0.006) were significantly associated with abnormal bowel habits. Each 5 additional missing teeth was associated with 7.1% higher odds of abnormal bowel habits (OR = 1.071, 95% CI: 1.027–1.117). Notably, constipation survived FDR correction among the 5 secondary outcomes (FDR-adjusted P = 0.032), with complete edentulism associated with 1.8-fold higher odds (OR = 1.766, 95% CI: 1.147–2.720, P = 0.013). This finding provides specific evidence for the tooth loss–constipation pathway and supports the masticatory dysfunction hypothesis. The association between tooth loss and abnormal bowel habits was robust across most sensitivity analyses, with modest attenuation noted after denture use adjustment in a restricted subset, and was particularly evident among adults aged <60 years and former smokers.

### 4.2 Comparison with Previous Studies

To our knowledge, this is the first study to examine the association between clinically examined tooth loss and bowel habits using the Bristol Stool Scale. A recent study by Ji et al.^[9]^ using NHANES 2005–2008 data investigated the relationship between self-reported oral health and BSS-assessed bowel habits, reporting associations between poor self-rated oral health and altered stool patterns. However, self-reported oral health is subject to recall bias and may not accurately reflect the objective degree of tooth loss. Our study extends these findings by using clinical dental examination data, which provides an objective, quantifiable measure of tooth loss. The use of the OHX component, in which trained dental examiners systematically assess each tooth, represents a significant methodological advance over self-reported measures. Our findings provide the first objective evidence supporting a link between clinically measured tooth loss and abnormal bowel habits.

### 4.3 Biological Mechanisms

Several biological mechanisms may underlie the observed association between tooth loss and abnormal bowel habits, particularly constipation. The primary proposed pathway involves masticatory dysfunction. Tooth loss, especially complete edentulism, substantially reduces chewing efficiency^[4]^. Impaired mastication has been associated with dietary modifications, including reduced intake of dietary fiber, fresh fruits, and vegetables, and increased consumption of soft, processed foods^[5; 18]^. Low dietary fiber intake is a well-established risk factor for constipation, as fiber increases stool bulk and accelerates colonic transit^[6; 19]^. A sensitivity analysis adjusting for dietary fiber intake showed that the association between tooth loss and abnormal bowel habits remained robust after fiber adjustment (P-for-trend: OR = 1.013, P = 0.006 vs. OR = 1.015, P = 0.001 without fiber; OR change: 0.2%), suggesting that while dietary fiber may partially mediate the tooth loss–bowel habit pathway, it does not fully explain the observed association. This finding implies that mechanisms beyond dietary fiber, such as the oral–gut microbiome axis, may play an important role.

An alternative or complementary mechanism involves the oral–gut microbiome axis. The oral cavity harbors the second-largest microbial community in the human body, and tooth loss and edentulism can significantly alter oral bacterial composition^[8]^. Oral bacteria are continuously swallowed and may contribute to the gut microbiome composition. Changes in oral microbial communities due to tooth loss may thus influence intestinal microbiota, potentially affecting gut motility and bowel function^[7; 20]^. Additionally, periodontal disease, which is a major cause of tooth loss, has been associated with systemic inflammation^[21]^, and chronic inflammation may affect gastrointestinal function through neuroimmune mechanisms^[22]^.

The finding that tooth loss is primarily associated with constipation rather than diarrhea is consistent with the masticatory dysfunction–dietary fiber hypothesis. Reduced fiber intake would be expected to slow colonic transit and promote constipation, without necessarily increasing diarrhea risk. The significant association between tooth loss and constipation that survived FDR correction (FDR-adjusted P = 0.032) provides the strongest evidence for this pathway. Although the 6–10 missing teeth group showed a significant association with diarrhea in categorical analysis (OR = 1.367, P = 0.048), the overall P-for-trend for diarrhea was not significant (P = 0.258), suggesting that this isolated finding may reflect a non-linear pattern or chance, and should be interpreted cautiously.

### 4.4 Dose–Response Relationship

The RCS analysis demonstrated a statistically significant nonlinear dose–response relationship (χ² = 11.84, df = 4, P = 0.019). The spline curve showed that the odds ratio remained near 1.0 from 0 to approximately 10 missing teeth, after which it began to rise gradually, with an apparent acceleration beyond approximately 14 missing teeth. At 28 missing teeth, the predicted OR was 1.452 (95% CI: 0.885–2.382). This threshold pattern is biologically plausible, as individuals with partial dentition may compensate through remaining teeth or prosthetic rehabilitation, whereas extensive tooth loss may exceed the compensatory capacity of the masticatory system.

The convergence of multiple analytical approaches—all yielding significant results—provides robust evidence for the dose–response relationship: the P-for-trend (P = 0.001), continuous variable analysis (P = 0.003), omnibus Wald test (P = 0.006), and RCS spline test (P = 0.019) all indicated a significant association. The continuous variable analysis estimated that each 5 additional missing teeth was associated with 7.1% higher odds of abnormal bowel habits (OR = 1.071, 95% CI: 1.027–1.117), providing a clinically interpretable measure of effect size.

The composite bowel outcome, integrating abnormal bowel habits, gas leakage, mucus leakage, and solid stool leakage, showed only borderline significance (P-for-trend = 0.052, omnibus P = 0.260). The high prevalence of the composite outcome (51.3%) reflects the dominance of the broad gas leakage definition (43.5% prevalence), which was not associated with tooth loss. In contrast, a sensitivity composite excluding gas leakage (17.8% prevalence) showed significant associations with tooth loss (P-for-trend = 0.010, continuous P = 0.008), with both the 6–10 missing teeth group (OR = 1.277, P = 0.040) and the edentulous group (OR = 1.425, P = 0.021) showing elevated odds. The persistence of the association after removing the non-associated gas leakage component demonstrates that the composite finding is not an artifact of the broad gas leakage definition and provides additional support for the tooth loss–bowel health relationship.

### 4.5 Subgroup Findings

The subgroup analysis revealed a borderline interaction between tooth loss and age (P-interaction = 0.080). Among participants aged <60 years, those with 11–28 missing teeth had significantly higher odds of abnormal bowel habits (OR = 1.486, 95% CI: 1.147–1.926, P = 0.004), whereas no association was observed among those aged ≥60 years (OR = 0.917, P = 0.516). This age-related pattern may reflect that tooth loss at a younger age is more likely to be associated with systemic health conditions or behavioral factors that also affect bowel function, whereas tooth loss in older adults is more normative and less indicative of underlying pathology. Alternatively, older adults may have had more time to adapt to dental prostheses, mitigating the functional consequences of tooth loss.

The association between tooth loss and abnormal bowel habits was significant among former smokers (OR = 1.651, 95% CI: 1.102–2.474, P = 0.019), while current smokers (OR = 1.223, P = 0.323) and never smokers (OR = 1.032, P = 0.813) showed no significant associations (P-interaction = 0.095). Smoking is a known risk factor for both periodontal disease and tooth loss^[23]^, and it may also independently affect gastrointestinal motility^[24]^. The significant association among former smokers may reflect that smoking cessation is often accompanied by other health behavior changes, or that former smokers represent a group with cumulative damage from smoking that persists after cessation. However, the interaction did not reach statistical significance at the 0.05 level and should be interpreted as suggestive rather than definitive.

Among non-diabetic participants, the association was significant (OR = 1.294, 95% CI: 1.051– 1.594, P = 0.019), while diabetic participants showed no significant association (OR = 1.115, P = 0.639; P-interaction = 0.174). No significant interactions were observed by sex (P-interaction = 0.220) or BMI (P-interaction = 0.289). Although elevated BMI has been shown to alter colonic sensory-motor function and transit^[17]^, the similar associations across BMI strata suggest that obesity-related physiological changes do not substantially modify the tooth loss–bowel habit relationship.

### 4.6 Sensitivity Analyses

The primary finding of an association between tooth loss and abnormal bowel habits was robust across most sensitivity analyses. The association remained significant after excluding participants with dental implants (6–10 teeth: OR = 1.311, P = 0.017; 28 teeth: OR = 1.567, P = 0.005), confirming that the result was not driven by implant-related confounding. When normal bowel habits were redefined as BSS 3–4 only, the association persisted (6–10 teeth: OR = 1.317, P = 0.016; 28 teeth: OR = 1.624, P = 0.004). The association between tooth loss and constipation was also robust when constipation was alternatively defined by bowel movement frequency <3 times/week (11–27 teeth: OR = 1.867, P = 0.029; 28 teeth: OR = 2.918, P < 0.001), providing strong support for the specificity of the tooth loss–constipation relationship.

A sensitivity analysis adjusting for dietary fiber intake was conducted using data from the NHANES 24-hour dietary recall files (DR1TOT). Among the 9,988 participants with dietary recall data (mean fiber intake: 16.1 g/day), the association between tooth loss and abnormal bowel habits remained robust: in the main analysis, the P-for-trend was OR = 1.015 (95% CI: 1.006–1.024, P = 0.001), and after additional adjustment for dietary fiber, the P-for-trend became OR = 1.013 (95% CI: 1.004–1.023, P = 0.006), representing only a 0.2% change in the odds ratio. The odds ratios for individual categories were also minimally affected (e.g., 28 missing teeth: OR = 1.545 → 1.482; 6–10 missing teeth: OR = 1.291 → 1.269). Dietary fiber itself was not significantly associated with abnormal bowel habits (OR = 0.990, 95% CI: 0.979–1.002, P = 0.089). The minimal attenuation of the tooth loss–bowel habit association after fiber adjustment suggests that dietary fiber is unlikely to be a major confounder of this relationship. However, because fiber intake may also lie on the causal pathway between tooth loss and bowel changes (i.e., as a mediator), the persistence of the association after adjustment indicates that mechanisms beyond reduced fiber intake—such as the oral–gut microbiome axis—likely contribute to the observed association.

An additional sensitivity analysis examined the potential confounding effect of denture use. Because denture use data (OHQ410 and OHQ430) were only collected in NHANES 2005–2008 and were unavailable for the 2009–2010 cycle, this analysis was restricted to a two-cycle subset (N = 6,424). After adjusting for denture use, the edentulism OR was modestly attenuated (from 1.464 to 1.316, a 10.1% reduction) and lost statistical significance (P = 0.033 to 0.203), while the P-for-trend remained essentially unchanged (0.120 to 0.123). Denture use itself was not an independent predictor of abnormal bowel habits (OR = 1.124, P = 0.401). The attenuation is consistent with denture use serving as a partial confounder—likely reflecting its strong correlation with age, socioeconomic status, and overall oral health deterioration—rather than a mediator on the causal pathway. However, the loss of statistical significance may also reflect reduced power in this smaller two-cycle subset (N = 6,424 vs. 9,988 in the primary analysis), as the P-for-trend in the same subset without denture adjustment was already non-significant (0.120). These findings suggest that while denture use may account for a portion of the observed association, it does not fully explain the dose–response relationship between tooth loss and abnormal bowel habits.

### 4.7 Strengths and Limitations

This study has several strengths. First, the use of clinical oral health examination data from trained dental examiners provides an objective, reliable measure of tooth loss, overcoming the limitations of self-reported measures used in previous studies. Second, the Bristol Stool Scale is a validated, standardized tool for assessing stool form, offering a more objective assessment of bowel habits than self-reported symptoms^[12]^. Third, the comprehensive NHANES dataset allowed for adjustment for a wide range of potential confounders, including demographic, socioeconomic, behavioral, and medical factors. Fourth, we examined multiple outcomes, including primary bowel habit categories and secondary fecal incontinence symptoms, providing a comprehensive assessment of the tooth loss–bowel health relationship. Fifth, we employed an a priori hierarchical testing framework^[13; 14]^, designating abnormal bowel habits as the primary outcome (analyzed via P-for-trend, exempt from multiplicity correction) and applying FDR correction only to the 5 secondary outcome comparisons, providing a transparent and statistically efficient approach to multiple comparisons. Sixth, the use of RCS allowed for detailed characterization of the dose–response relationship, which achieved statistical significance (P = 0.019). Seventh, the large sample size (N = 9,988), including 753 edentulous participants, provided adequate statistical power across all tooth loss categories. Finally, the use of a composite bowel outcome provided a sensitive measure of the overall bowel symptom burden associated with tooth loss, and a sensitivity composite excluding gas leakage confirmed that this finding was not driven by the broad gas leakage definition.

Several limitations should be acknowledged. First, the cross-sectional design precludes causal inference. Although we hypothesize that tooth loss leads to altered bowel habits through masticatory dysfunction and dietary changes, reverse causality cannot be excluded—individuals with gastrointestinal disorders may alter their diet in ways that promote dental caries and tooth loss, or conditions such as gastroesophageal reflux disease may directly damage tooth enamel and subsequently lead to tooth loss. Second, although a sensitivity analysis adjusting for dietary fiber intake was conducted across all three survey cycles (Section 4.6), dietary fiber was assessed using a single 24-hour dietary recall (Day 1), which may not reflect habitual intake. Other dietary factors such as fluid intake, fat consumption, and overall diet quality were not controlled for and may contribute to residual confounding. Third, medication use was not controlled for. Numerous medications, including laxatives, antidiarrheals, opioids, anticholinergics, calcium channel blockers, and antidepressants, can affect bowel habits and may confound the observed associations. Fourth, the BSS assessment in NHANES reflects a single measurement, which may not capture the day-to-day variability in stool form. Fifth, the high prevalence of “any” gas leakage (43.5% when using the broad definition of responses 1–5) is consistent with the known high prevalence of mild fecal incontinence in the general population, particularly gas incontinence, which is the most common and least severe form. Importantly, gas leakage was not associated with tooth loss in regression analyses, and the composite outcome including gas leakage showed only borderline significance, whereas the sensitivity composite excluding gas leakage was significant. This indicates that the high prevalence of gas leakage reflects the common nature of mild symptoms rather than a true association with dental status. Sixth, physical activity was assessed using different questionnaires across survey cycles (GPAQ in 2007–2010; PAQ180 self-rated comparison in 2005–2006), and the binary active/inactive classification may not fully capture the complexity of physical activity; this cross-cycle inconsistency could introduce measurement noise. Seventh, complete-case analysis was used to handle missing covariate data, which assumes data are missing completely at random (MCAR); if missingness is related to both tooth loss and bowel habits, selection bias may be introduced. Eighth, although we adjusted for numerous covariates, residual confounding by unmeasured factors (e.g., gastrointestinal diagnoses, Helicobacter pylori infection, socioeconomic factors not captured by PIR) cannot be excluded. Ninth, although we employed a hierarchical testing framework to address multiplicity, the secondary and supplementary analyses beyond constipation remain exploratory and require replication. While constipation survived FDR correction, the other secondary outcomes (diarrhea, gas leakage, mucus leakage, solid stool leakage) did not reach significance and should be interpreted as hypothesis-generating. Tenth, the effect sizes observed in this study are modest (ORs of 1.3–1.5 for the primary outcome), which is consistent with the multifactorial nature of bowel habits but limits the clinical significance of the findings at the individual level. Eleventh, the wide confidence interval for the edentulism–constipation association (95% CI: 1.147–2.720) reflects the limited number of constipation events among edentulous participants (n = 72), which constrained statistical power for this specific subgroup analysis despite the overall significant trend (P-for-trend = 0.006).

Finally, the study population was limited to adults aged ≥20 years who participated in NHANES 2005–2010, which may limit generalizability to other populations. Additionally, denture use data were only available for the 2005–2008 cycles, precluding adjustment for this variable in the full sample; the sensitivity analysis on the available subset suggested partial confounding by denture use.

### 4.8 Clinical Implications

Our findings suggest that tooth loss is a risk marker for abnormal bowel habits and broader bowel symptoms, particularly among individuals with extensive tooth loss. The significant association with constipation, which survived FDR correction, supports the hypothesis that masticatory dysfunction may contribute to altered bowel habits through reduced dietary fiber intake and other mechanisms. Clinicians caring for edentulous patients should be aware of this potential association and may consider screening for bowel symptoms. From a public health perspective, the findings underscore the importance of maintaining natural dentition for overall health, including digestive function. Preventive dental care and timely prosthodontic rehabilitation may help mitigate the dietary consequences of tooth loss and potentially reduce the risk of associated bowel dysfunction. However, prospective studies with longitudinal dietary assessments are needed to confirm these findings and elucidate the underlying mechanisms.

## 5. Conclusions

In this nationally representative cross-sectional study of US adults, tooth loss was significantly associated with abnormal bowel habits in the a priori primary analysis (P-for-trend = 0.001), corroborated by the omnibus Wald test (P = 0.006), continuous variable analysis (P = 0.003), and restricted cubic spline analysis (P = 0.019). Each 5 additional missing teeth was associated with 7.1% higher odds of abnormal bowel habits. Constipation survived FDR correction among secondary outcomes (FDR-adjusted P = 0.032), providing specific evidence for the tooth loss– constipation pathway. The association was robust across most sensitivity analyses, with modest attenuation noted after denture use adjustment in a restricted subset. These findings support the hypothesis that masticatory dysfunction associated with tooth loss may contribute to altered bowel habits. Longitudinal studies with dietary assessment are needed to confirm causality and elucidate the underlying mechanisms.

## Materials

The following are available online at [website]: Table S1: STROBE checklist for cross-sectional studies.

## Author Contributions

HC contributed to the study design, data analysis, interpretation of results, and manuscript drafting. JZ contributed to the study concept, critical revision of the manuscript, and supervision of the research. All authors read and approved the final manuscript.

## Data Availability Statement

NHANES data are publicly available at https://www.cdc.gov/nchs/nhanes/ (accessed on [date]).

## Acknowledgments

The authors thank the NCHS for making NHANES data publicly available.

## Ethics Statement

The NHANES protocol was approved by the National Center for Health Statistics (NCHS) Research Ethics Review Board. All participants provided written informed consent. This secondary analysis of de-identified, publicly available data did not require additional institutional review board approval.

## References

[1] N.J. Kassebaum, E. Bernabé, M. Dahiya, B. Bhandari, C.J. Murray, and W. Marcenes, Global Burden of Severe Tooth Loss: A Systematic Review and Meta-analysis. J Dent Res 93 (2014) 20s–28s.

[2] P.E. Petersen, D. Bourgeois, H. Ogawa, S. Estupinan-Day, and C. Ndiaye, The global burden of oral diseases and risks to oral health. Bull World Health Organ 83 (2005) 661–9.

[3] M.S. Tonetti, S. Jepsen, L. Jin, and J. Otomo-Corgel, Impact of the global burden of periodontal diseases on health, nutrition and wellbeing of mankind: A call for global action. J Clin Periodontol 44 (2017) 456–462.

[4] A. Sheiham, J.G. Steele, W. Marcenes, C. Lowe, S. Finch, C.J. Bates, A. Prentice, and A.W. Walls, The relationship among dental status, nutrient intake, and nutritional status in older people. J Dent Res 80 (2001) 408–13.

[5] Y. Zhu, and J.H. Hollis, Tooth loss and its association with dietary intake and diet quality in American adults. J Dent 42 (2014) 1428–35.

[6] S.A. Müller-Lissner, M.A. Kamm, C. Scarpignato, and A. Wald, Myths and misconceptions about chronic constipation. Am J Gastroenterol 100 (2005) 232–42.

[7] T.S. Schmidt, M.R. Hayward, L.P. Coelho, S.S. Li, P.I. Costea, A.Y. Voigt, J. Wirbel, O.M. Maistrenko, R.J. Alves, E. Bergsten, C. de Beaufort, I. Sobhani, A. Heintz-Buschart, S. Sunagawa, G. Zeller, P. Wilmes, and P. Bork, Extensive transmission of microbes along the gastrointestinal tract. Elife 8 (2019).

[8] B.J. Kunath, C. De Rudder, C.C. Laczny, E. Letellier, and P. Wilmes, The oral-gut microbiome axis in health and disease. Nat Rev Microbiol 22 (2024) 791–805.

[9] Z. Ji, J. Mei, Y. Li, Z. Wang, Z. Guo, and L. Miao, Association between oral health and bowel habits: a cross-sectional study. BMC Public Health 25 (2025) 1462.

[10] G. Zipf, M. Chiappa, K.S. Porter, Y. Ostchega, B.G. Lewis, and J. Dostal, National health and nutrition examination survey: plan and operations, 1999-2010. Vital Health Stat 1 (2013) 1–37.

[11] B.A. Dye, J. Afful, G. Thornton-Evans, and T. Iafolla, Overview and quality assurance for the oral health component of the National Health and Nutrition Examination Survey (NHANES), 2011-2014. BMC Oral Health 19 (2019) 95.

[12] S.J. Lewis, and K.W. Heaton, Stool form scale as a useful guide to intestinal transit time. Scand J Gastroenterol 32 (1997) 920–4.

[13] K.J. Rothman, No adjustments are needed for multiple comparisons. Epidemiology 1 (1990) 43–6.

[14] D.A. Savitz, and A.F. Olshan, Multiple comparisons and related issues in the interpretation of epidemiologic data. Am J Epidemiol 142 (1995) 904–8.

[15] R. Bender, and S. Lange, Adjusting for multiple testing--when and how? J Clin Epidemiol 54 (2001) 343–9.

[16] T. Lumley, Analysis of complex survey samples. Journal of Statistical Software 8 (2004) 1–19.

[17] S. Delgado-Aros, M. Camilleri, M.A. Garcia, D. Burton, and I. Busciglio, High body mass alters colonic sensory-motor function and transit in humans. Am J Physiol Gastrointest Liver Physiol 295 (2008) G382–8.

[18] A.W. Walls, J.G. Steele, A. Sheiham, W. Marcenes, and P.J. Moynihan, Oral health and nutrition in older people. J Public Health Dent 60 (2000) 304–7.

[19] J.W. McRorie, Jr., and N.M. McKeown, Understanding the Physics of Functional Fibers in the Gastrointestinal Tract: An Evidence-Based Approach to Resolving Enduring Misconceptions about Insoluble and Soluble Fiber. J Acad Nutr Diet 117 (2017) 251– 264.

20. K. Atarashi, W. Suda, C. Luo, T. Kawaguchi, I. Motoo, S. Narushima, Y. Kiguchi, K. Yasuma, E. Watanabe, T. Tanoue, C.A. Thaiss, M. Sato, K. Toyooka, H.S. Said, H. Yamagami, S.A. Rice, D. Gevers, R.C. Johnson, J.A. Segre, K. Chen, J.K. Kolls, E. Elinav, H. Morita, R.J. Xavier, M. Hattori, and K. Honda, Ectopic colonization of oral bacteria in the intestine drives T(H)1 cell induction and inflammation. Science 358 (2017) 359–365.

[21] B.L. Pihlstrom, B.S. Michalowicz, and N.W. Johnson, Periodontal diseases. Lancet 366 (2005) 1809–20.

[22] J.B. Furness, The enteric nervous system and neurogastroenterology. Nat Rev Gastroenterol Hepatol 9 (2012) 286–94.

[23] S. Warnakulasuriya, T. Dietrich, M.M. Bornstein, E. Casals Peidró, P.M. Preshaw, C. Walter, J.L. Wennström, and J. Bergström, Oral health risks of tobacco use and effects of cessation. Int Dent J 60 (2010) 7–30.

[24] W.M. McDonnell, and C. Owyang, Effects of smoking on interdigestive gastrointestinal motility. Dig Dis Sci 40 (1995) 2608–13.

